# Democratizing Virtual Patient Case Creation: A Proof-of-concept Technical Framework for Clinicians

**DOI:** 10.1101/2024.10.25.24315696

**Authors:** Nikolaos Tsaftaridis, Ioannis Koulas, Stefanos Zafeiropoulos, Veauthyelau Saint-Joy, Marwa Ilali, Michel Ibrahim, Taina Brice, Norrisa Haynes

**Affiliations:** Feinstein Institutes for Medical Research, Northwell Health; Global MedEd Network; Department of Cardiology, University Hospital of Zurich; Department of Family Medicine, McGill University; Cardiovascular Medicine, ChenMed; Yale School of Medicine; Yale Institute for Global Health

**Keywords:** Twine, Virtual Patients, Case-based Learning, Echocardiography, Global Health

## Abstract

**Objective:** Virtual patient cases are a scalable and engaging tool for training medical professionals. Strategies and frameworks for their implementation in teaching and training settings are few, technically complicated and/or expensive. We developed and evaluated open source and free virtual patient cases to test knowledge acquisition during an echocardiography training program for internal medicine trainees in Haiti. The objective of this paper is to describe the technical aspects of the GMENEcho virtual patient cases implementation and motivate similar work by resource-constrained teams.

**Methods:** We used an open source engine for text-based games (Twine) since it provides the necessary interaction mechanics and is usable out-of-the-box. The case code was written in SugarCube 2.30.0 notation and the tweego-generated .html file was hosted on Github Pages for continuous integration and deployment, making iterations by the clinical team seamless. Data from completed tests were reported back via email through a third party integration.

**Results:** The technical work was completed in two weeks by a team member with a clinical background and minimal computer programming experience. The virtual patient cases were deployed for a pretest (November 2023) and a second time unaltered for a posttest (June 2024) after the interim hands-on and theoretical training had been completed. Qualitative feedback was positive or neutral. The overall score in the posttest was significantly higher with a large effect size (mean absolute improvement 15.26%, p < 0.001; Cohen’s d: 1.398), similarly to the diagnostic score (mean absolute difference 16.09%, p < 0.001; Cohen’s d: 1.402). Management performance missed statistical significance by a small margin. The System Usability Scale (SUS) score was 74.6 (“Excellent”).There was reduced inter-trainee variability across metrics in the posttest, including the SUS score.

**Discussion:** This proof-of-concept methodology can be applied to create clinical patient cases for use within a class or a clinical training setting, through a friendly graphical user interface. A more complex software stack can allow for remote or larger scale implementations with additional features.

**Conclusion:** The rapid development time and positive qualitative and quantitative feedback highlight the potential of this approach for clinical education in resource-constrained settings. It can serve as a template for more streamlined adaptations of case-based learning in diverse healthcare settings.

## 1 Introduction

Case-based learning is an adjunct teaching modality that aims to connect theory and practice by engaging the trainee in a conversational and active process of learning through the provision of simulated (in text or otherwise) patient cases, along with rapid and individualized feedback.^1,2^

In the context of medical education several gamified educational apps have been created that aim to provide time-efficient learning to trainees (McCoy, Lewis and Dalton, 2016). ^3^ Virtual patient scenarios seem to be an engaging method to deliver clinical vignettes relevant to a physician’s duties. ^4^ However, there are limited software options that can be deployed independently for the creation and delivery of such scenarios. When open source options are available, such as OpenLabyrinth, they require significant digital infrastructure and/or programming ability. ^5^

As a digital learning modality requiring relevant infrastructure and expertise, virtual patient scenarios have been less readily available in resource-constrained settings.^6^ Lack of infrastructure and difficulties with cultural and educational adaptation are examples of the typical barriers educators face in the implementation of simulation-based didactics in low income countries. ^7^

Additionally, most available online clinical cases are targeted at specific exams (i.e. USMLE, COMLEX, board licensing exams, etc.), with high costs to build, deploy and access. These platforms are not cognizant of local conditions and cultural characteristics except for the state of the art in high-resource academic settings. They are also not modifiable, extendable or replicable, limiting the potential for dissemination in other contexts.

Given the socioeconomic and political challenges in Haiti, there is a great need to establish resilient cardiology training modalities and programs.^8^ As part of the “Focused Echo InTervention Package” (FETIP) project, we created virtual patient cases that would test the echocardiography knowledge of internal medicine trainees in Haiti in a manner responsive to local needs and realities. The purpose of this paper is to describe the technical implementation of our virtual patient cases in a manner approachable to interested parties with minimal technical expertise and encourage case-based learning in all settings.

## 2 Methods

The GMENEcho digital patient cases were intended to bookend the FETIP program as an assessment module. The FETIP project was a 6-month-long educational intervention implemented in the University Hospital of La Paix, one of the four University Hospitals in Haiti. The technological infrastructure includes computers and smartphones with internet access, allowing for digital educational interventions. There are, however, significant challenges, including electricity interruptions and political instability, which can affect the consistency of access to these technologies. Additionally, there is a lack of hands-on programs for technical skill development with regards to echocardiography training.

This virtual patient case system was developed to serve as both an assessment tool for the GMENEcho training program and a potential standalone educational resource for cardiovascular medicine in resource-constrained settings.

To quantitatively measure the impact of our cases, we calculated absolute improvements and standardized differences between the pretest and posttest scores. Three scores were calculated as main outcomes by evaluating performance across quizzes and scenario-based actions for all four cases: overall performance, diagnostics and management scores. Interpretation of echocardiographic images was presented to the trainees as quiz questions and incorporated into diagnostic scores. Details on the contribution of the case questions and choices to the scores are available in Figure 4 of the Supplement. Statistical significance testing was performed using paired samples t-tests, given the small number of participants (<20). Effect size assessment was conducted using Cohen’s d. We used the rule of thumb thresholds set for small (0.1 to <0.30), moderate (0.3 to <0.5), and large (≥0.5) effect sizes. Statistical analysis was conducted and graphs were created using Jamovi Version 2.5.

### 2.1 Rationale

The development of the cases was a multidisciplinary effort involving clinical and research cardiologists along with medical researchers from Haiti, Greece and the United States. The clinical content of the cases was created with the direction of on-the-ground physicians to address pain-points in cardiology care in Haiti.

The cases tied together clinical presentation, workup, echocardiographic imaging acquisition, interpretation and clinical management in a choose-your-own-adventure format, where choices affected patient outcomes and the performance was graded qualitatively on scale from insufficient to optimal.

The content was culturally adapted, with scenarios tailored to the Haitian healthcare context and incorporation of locally available resources and treatment options. This adaptation was crucial to ensure the relevance and applicability of the cases to the target audience. The clinical details of the scenarios are further discussed in a separate manuscript (in preparation).

### 2.2 Design

Given the limited budget, to enhance immersion and engagement, we used stable diffusion-generated images depicting human interactions between patients and medical staff and conveying positive or negative sentiment depending on how the trainee was doing in the case.^9^ The prompts were adapted to generate culturally appropriate images and to minimize the bias inherent in the image generating models. Examples of the prompts and resulting images are shown in Figure 1.

**Figure 1.**
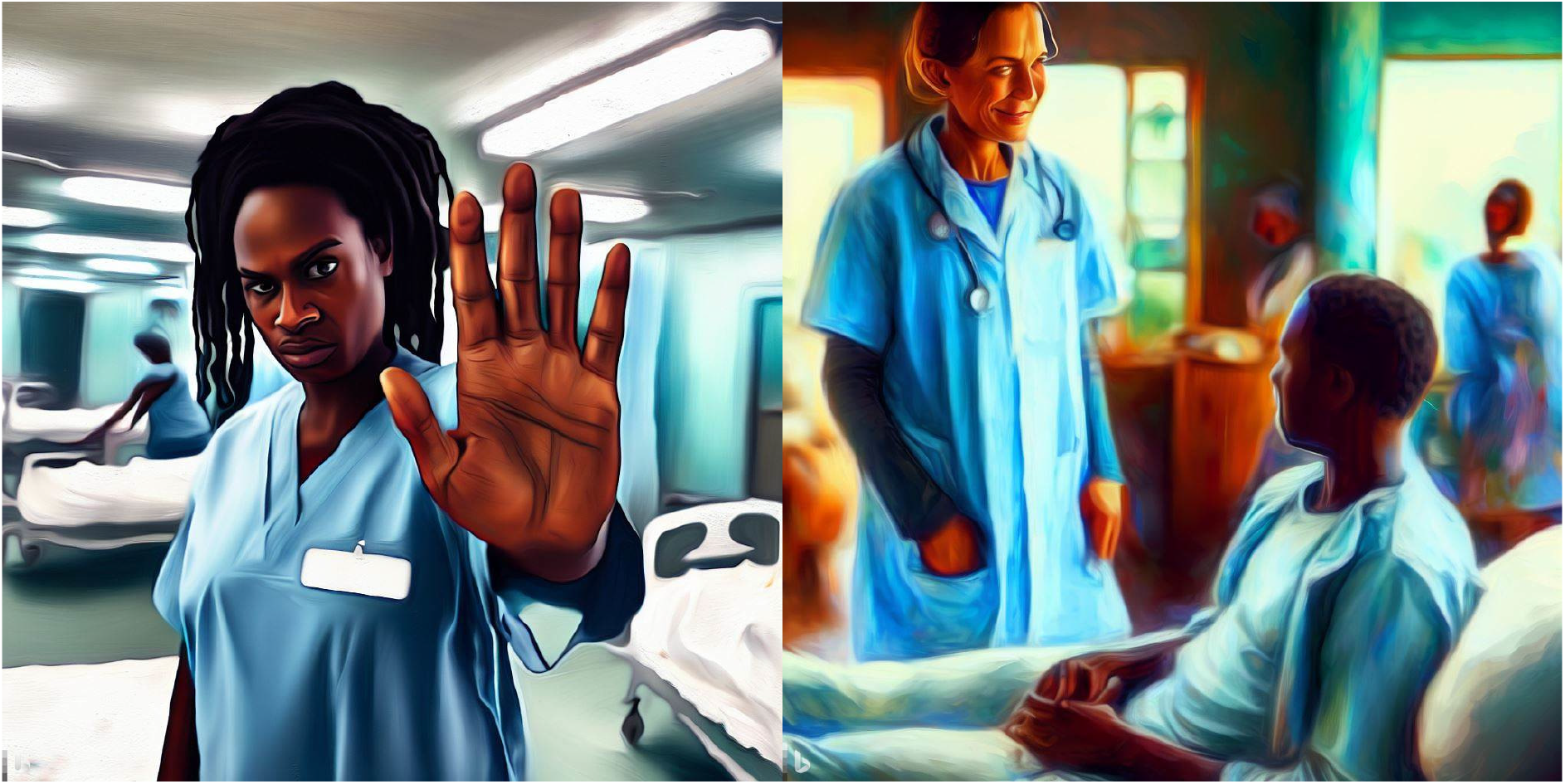
Examples of AI-generated images used as illustrations in the GMENEcho Project. Prompts used were “A realistic digital painting of an African nurse in a hospital ward looking at the camera doing the stop sign frightened, precise, high resolution” (left) and “A realistic digital painting of a doctor in Haiti smiling at a patient who is talking to him from his bed, detailed, vibrant colors, nurses in the background, high resolution” (right). Notice doctor on the right is presented as white, in an example of bias inherent to the image generating model.

Multimedia including ECGs, heart sound audio, chest X-rays, transthoracic and transesophageal echo recordings were selected from open educational repositories to fit each specific case.^10–13^

### 2.3 Development

Development began in March 2023 and concluded in September 2023. The core technical work was completed in a period of two weeks, taking about 15 hours of work and the rest of the time was dedicated to the iterative improvement of the clinical case content.

After developing the academic content for each case, lessons learned were taken into consideration to improve the implementation of the next one. The initial design of the interaction model featured multiple-choice questions allowing for a single selection. This was later evolved to allow trainees to select multiple or no treatment options, mirroring real-life clinical decision-making processes. Many of the nodes in each scenario were interconnected so that users could select between different diagnostic modalities and then returning back to the main node where they would decide the treatment based on their assessment. In the case of an erroneous choice, the script offered advice to guide the user into the correct choice, in the example of a nurse reminding the user that their selection is wrong and prompting them to reconsider it. These intermediate nodes were only available to the user if they had selected a wrong choice and aimed to provide immediate feedback and redirect the user to the right steps in order to complete the scenario.

Each case was structured with an interactive progression, beginning with a patient presentation and vital signs. Users were required to navigate through various diagnostic choices, including physical exam, EKG, chest X-ray, labs, and echocardiography. The cases incorporated multimedia elements such as audio clips of heart sounds, EKG images, chest X-ray images, and echocardiogram videos and still images. Based on their diagnostic findings, users were then prompted to make treatment decisions. The cases also included follow-up management and patient outcomes to provide a comprehensive learning experience.

A scoring system was implemented to track user performance in diagnosis, treatment, and follow-up management. Initial implementations incorporated both choice tracking and realtime scoring. However, due to complexity concerns, later iterations focused solely on choice tracking, deferring scoring to post-hoc data analysis.

Given that the cases were originally created in English, translation into French and Haitian Creole was undertaken by team members fluent in French and local Haitian Creole idioms, aided via automatic translation of the English text through free online translation services. This significantly reduced the time necessary to deploy the French version of the cases.

In order to evaluate this effort we aimed to quantify how the end users felt when using the scenarios and how well they did in their medical decision making throughout the scenarios. The System Usability Scale (SUS) questionnaire (Brooke, 1996) was deployed after all cases were completed to collect information about the usability of the virtual patients platform, with responses graded on a 5-point Likert scale from 1 = completely disagree to 5 = completely agree. This is an industry-standard tool that can provide an overall score based on the end-user responses as follows: <25 = “Worst Imaginable”, 25.1–51.6 = “Poor”, 51.7–62.6 = “OK/Fair”, 62.7–72.5 = “Good”, 72.6–84 = “Excellent”, >84.1 = “Best Imaginable”.

The details and results of the academic scoring of the scenarios are described in more detail in the clinically-directed manuscript of the main GMENEcho study (in preparation).

### 2.4 Technical Implementation

Each virtual patient case was scripted using Twine’s SugarCube notation (2.30.0) and saved as an individual source file. Additional files were created to handle user identification, usability testing, and score reporting, as well as CSS style customization and JavaScript for custom engine functionality. All these components were compiled into a browser-readable static website format using the command line tool Tweego, with continuous deployment during development and updates managed through GitHub Actions. The static website files generated by Tweego were then hosted on GitHub Pages under a custom domain. This allowed us to deliver a complete web page in the form of a single HTML file, which could be downloaded to the trainee’s device in one go, just by visiting our webpage URL. This setup avoided the need for expensive server hosting and minimized server-client interaction.

Trainees were identified by entering a unique password of their choosing to start solving the cases. This was also used to track them between the pre- and posttest. All trainee actions in a static website are logged and managed client-side, using pre-generated standard HTML. User feedback and the results of each session were collected via an HTML form submission, through Static Forms—a service forwarding the data to the developers’ email address.^14^ This method was deemed appropriate due to the low expected volume of participants in this pilot. A graphical representation of this process can be seen in Figure 2.

**Figure 2.**
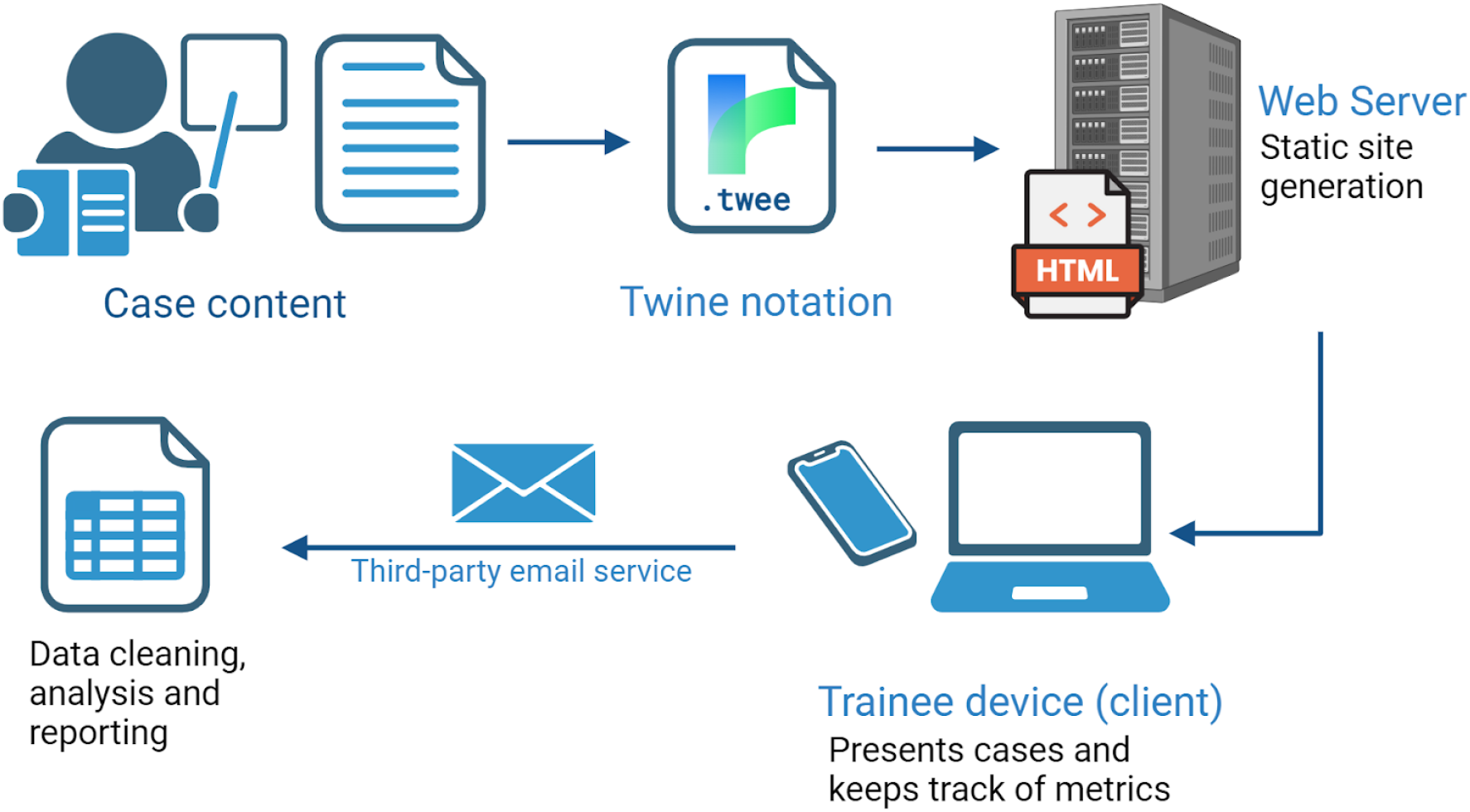
GMENEcho Technical Workflow: After case content creation, the scenarios were translated into Twine notation and uploaded as a static website (single .html file) on a free hosting server. After case completion, results were reported back to the research team via a thirdparty email service. The data were then imported, cleaned and analyzed manually. Created in BioRender. Koulas, I. (2024) BioRender.com/g96n129

**Figure 3.**
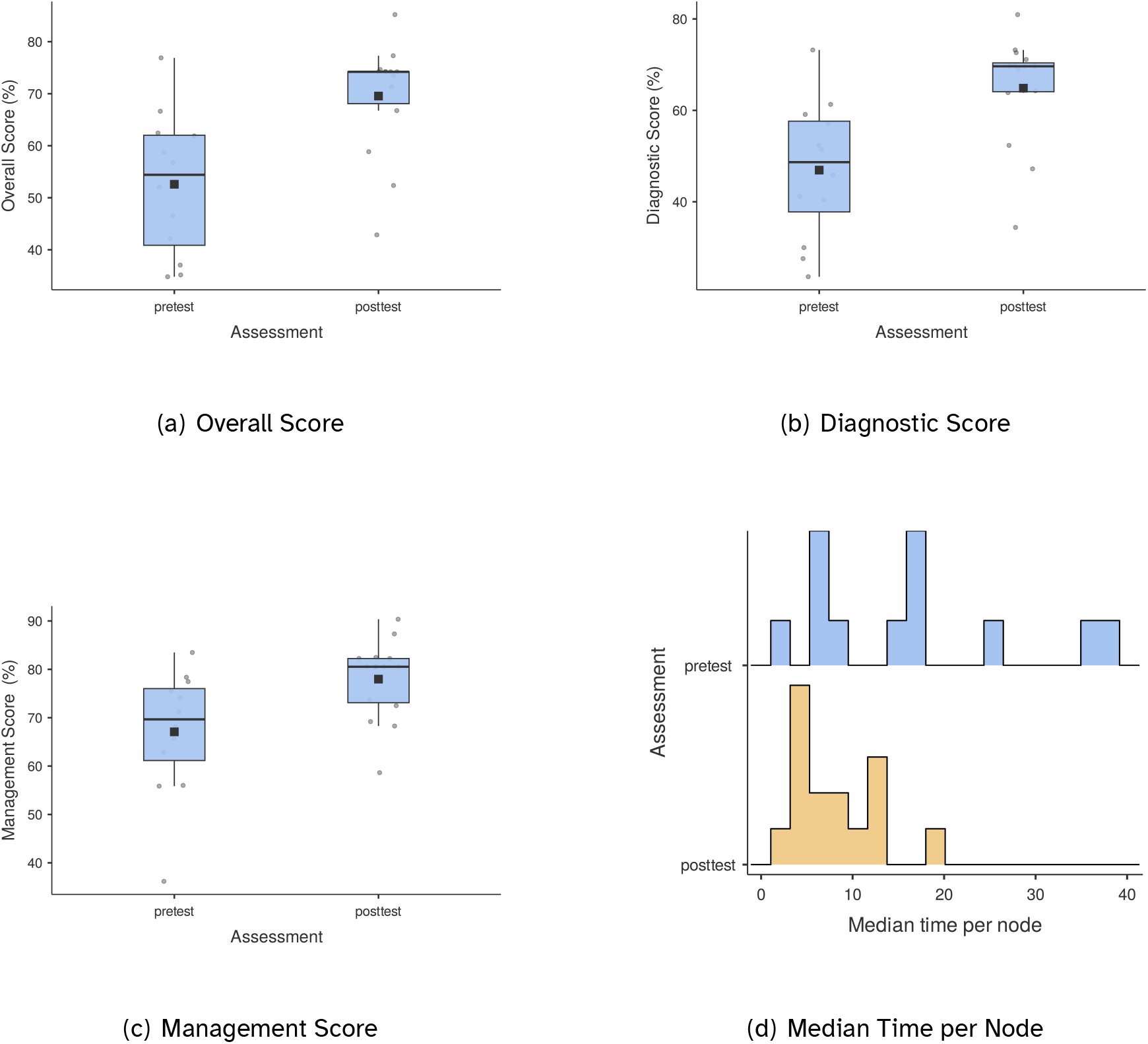
Average overall score, diagnostic score and management score box plots. Median time per node/step through all cases per trainee (histogram).

**Figure 4.**
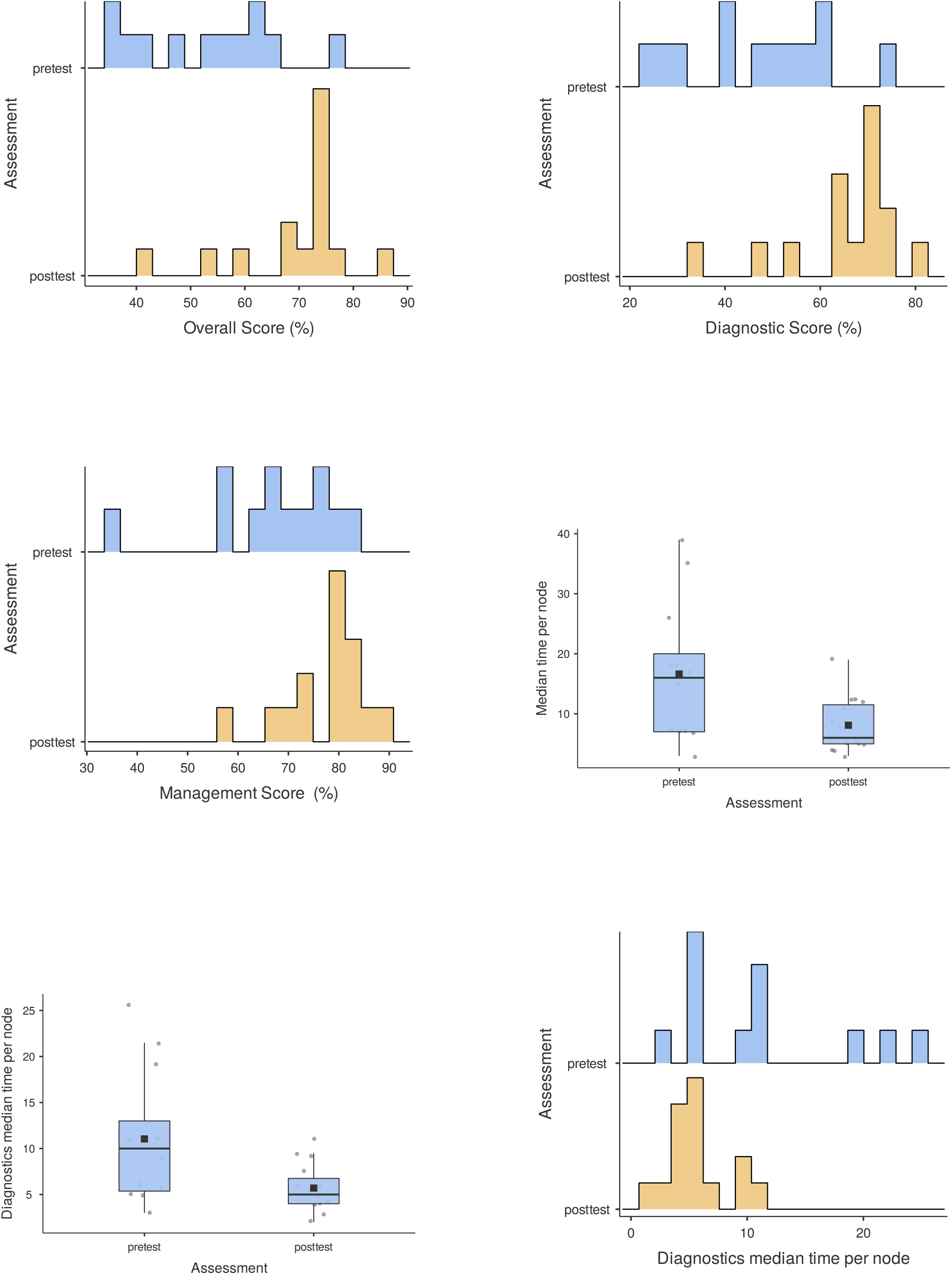
Complimentary histograms and box plots for Figure 3

**Figure 5.**
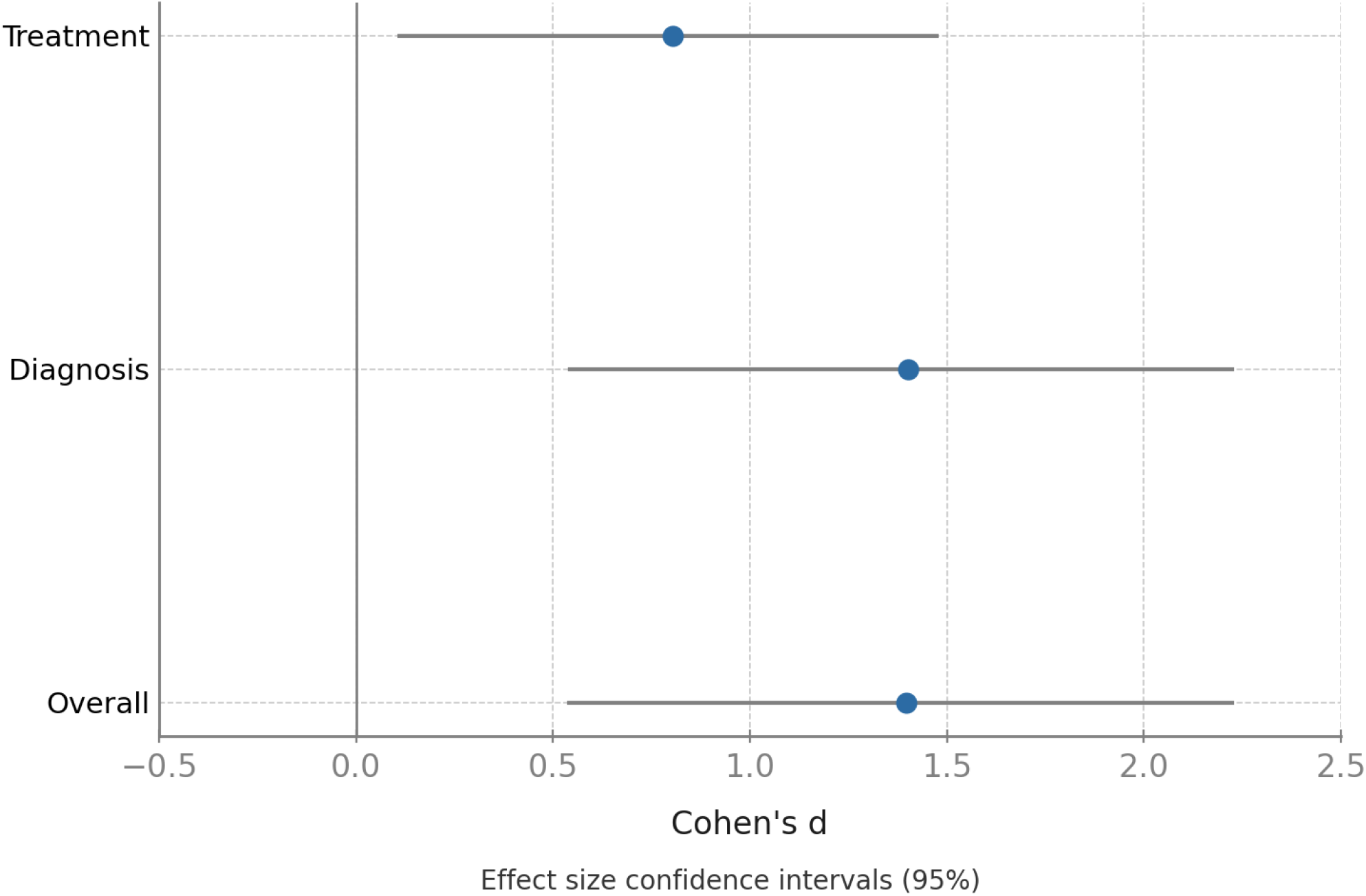
95% confidence intervals for the three comparisons.

**Figure 6.**
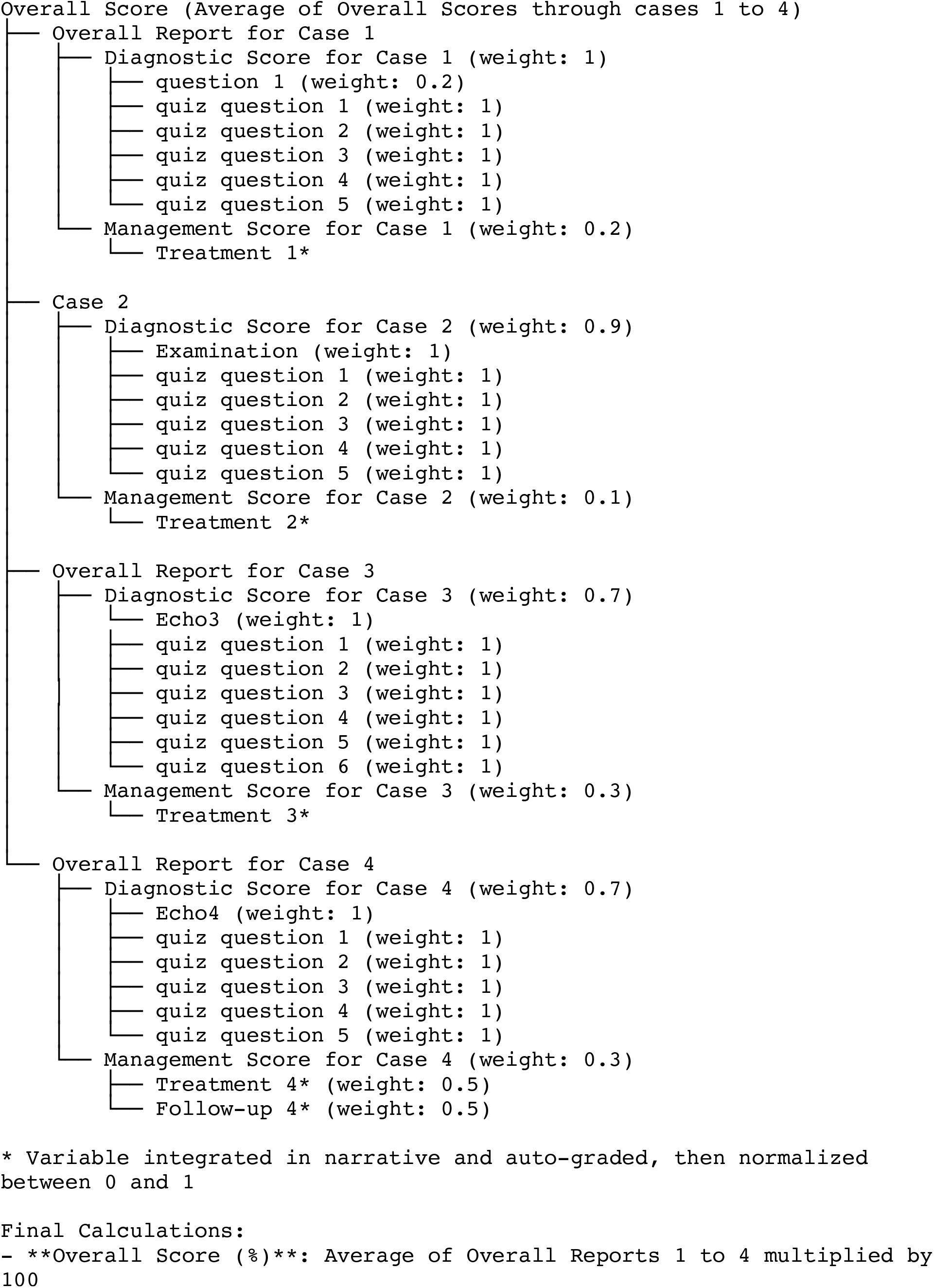
Schematic representation of weights and contributions of assessment data to the reported scores.

No sensitive patient or trainee information was collected. Prototyping was completed without explicit cybersecurity considerations other than the security inherent in the hosting platform.

For technical details, please refer to the project’s code repository. ^15^

## 3 Results

The trainees were able to fully access the cases via web browsers either in their mobile devices or their personal computers. The material was delivered independently of client screen dimensions ensuring minimal issues with responsiveness. No data were received or sent from and to the server once the scenarios were loaded.

Out of a total of 16 trainees, 12 completed the pretest and 15 completed the posttest (Table 1). There was a statistically significant improvement in overall scores and diagnostic scores between the pretest and posttest, while management scores closely missed statistical significance (Table 2). Large effect sizes were observed for all three scores.

**Table 1:**
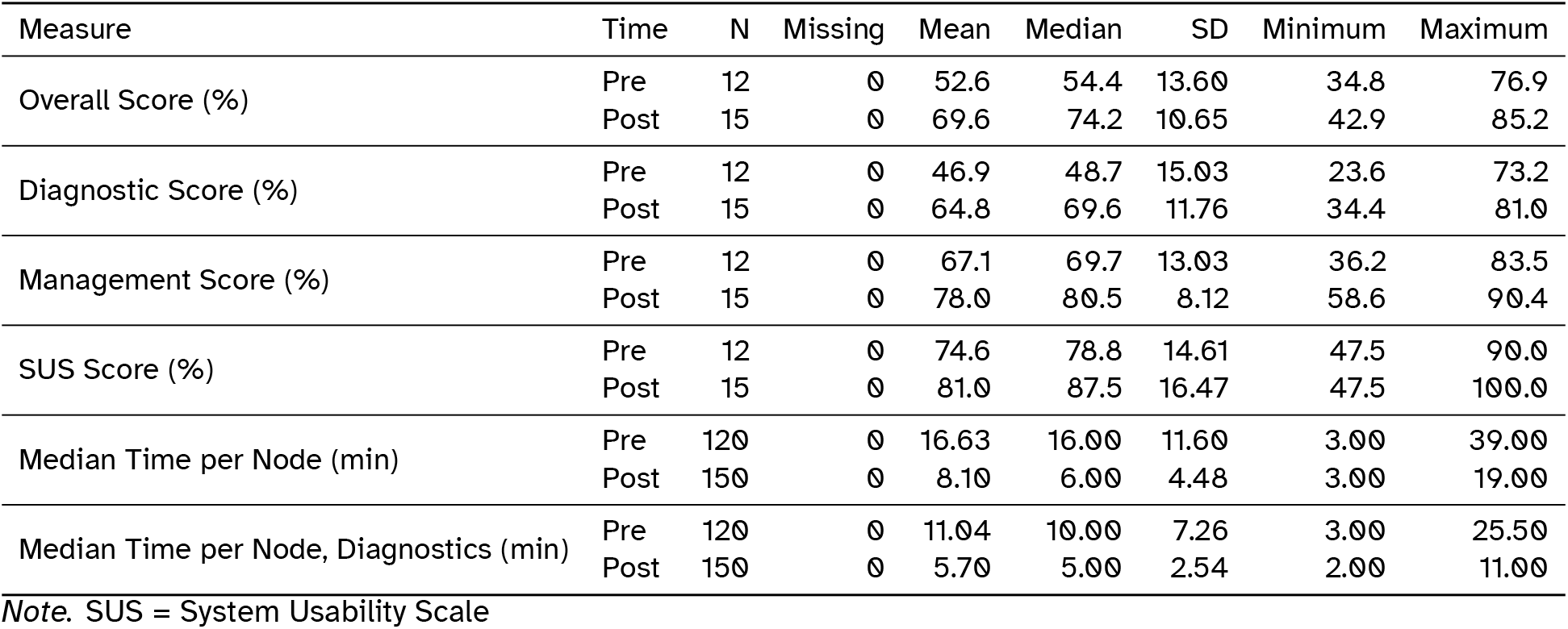
Descriptive Statistics for the Pretest and Posttest Assessments.

**Table 2:** Paired Samples t-test Results.

|  |  |  | statistic | df | p | Mean diff | Cohen's d |
| --- | --- | --- | --- | --- | --- | --- | --- |
| Overall Score Post | Overall Score Pre | Student's t | 4.64 | 10.0 | <0.001 | 15.26 | 1.398 |
| Diagnostic Report Post | Diagnostic Report Pre | Student's t | 4.65 | 10.0 | <0.001 | 16.09 | 1.402 |
| Management Score Post | Management Score Pre | Student's t | 2.67 | 10.0 | 0.012 | 9.84 | 0.805 |

Note. $H_a: \mu_{\text{Measure 1} - \text{Measure 2}} > 0$ ; df = degrees of freedom; Mean diff = Mean difference, (%)

**Table 3:** Shapiro-Wilk Test Results for Normality.

|  |  |  | W | p |
| --- | --- | --- | --- | --- |
| Overall Score Post | - | Overall Score Pre | 0.954 | 0.691 |
| Diagnostic Report Post | - | Diagnostic Report Pre | 0.959 | 0.761 |
| Management Score Post | - | Management Score Pre | 0.917 | 0.296 |

Note. A low p-value suggests a violation of the assumption of normality

The overall score in the posttest was significantly higher than the pretest, (mean absolute score increase 15.26%, p < 0.001) and a large effect size (Cohen’s d: 1.398). The diagnostics score reports showed a similarly significant and large improvement (mean absolute difference 16.09%, p < 0.001 and Cohen’s d: 1.402). Management scores also improved, though statistical significance was not reached (mean absolute difference 0.98, *p* : 0.012, Cohen’s d: 0.805). Confidence intervals for the effect size are included in the supplement.

In terms of time efficiency, median time per page decreased from 16.63 seconds to 8.10 seconds between the two assessments, with diagnostic-specific time decreasing from 11.04 to 5.7 minutes.There also was reduced variability across metrics in the posttest, including the SUS score.

Qualitative feedback from participants indicated a generally positive reception of the virtual patient cases. Trainees found the cases useful for understanding key concepts, prioritizing diagnostic procedures, and summarizing clinical scenarios. The cases were perceived as realistic, with participants noting that they reflected common clinical presentations seen in their hospital. Some suggestions for improvement included gradually increasing the difficulty of echocardiographic interpretations and highlighting specific diagnostic criteria for each clinical vignette.

Participants also appreciated the inclusion of culturally relevant terms and patient-physician interactions, though they recommended addressing issues like treatment interruptions due to financial constraints.

## 4 Discussion

This study presents a low-cost framework for the development and deployment of virtual patient cases tailored specifically to enhance echocardiography training in resource-limited settings. Our approach leverages open-source technology to provide scalable and cost-effective educational tools.

### 4.1 Outcomes

In terms of the results, three general knowledge domains were assessed: cardiovascular disease diagnosis, cardiovascular disease management, and an aggregate score combining both. The questions were primarily weighted towards diagnosis, as the curriculum emphasized the ability to diagnose and characterize different types of heart failure based on clinical presentation and echocardiography. This emphasis may have underpowered the management component of the assessment, which likely explains an improvement in management post-test scores and a large effect size that did not reach statistical significance.

The results, which showed statistically significant large effect sizes in two of the three main metrics (overall score and diagnostic scores), highlight the feasibility and practicality of using custom patient cases to evaluate knowledge improvement over time as part of a broader educational intervention in resource-constrained settings. The same questions were included in both the pre- and post-tests, though presented in different orders with varied multiple-choice options. Additionally, the six-month gap between pre- and post-tests likely minimized recall bias. Some participants submitted perfect scores (100%) on the post-test SUS evaluation, and there was an observed increase in both score improvement and efficiency (reduced time to completion).

### 4.2 Lessons Learned

In terms of methodology, at the outset, we defined precise objectives aimed at evaluating the efficacy of the echocardiography training program through contextual clinical scenarios. Unlike traditional teaching and testing tools, our virtual patient cases were designed to simulate local clinical environments, fostering comprehensive clinical skill evaluation.

We utilized an iterative integration and deployment pipeline which enabled us to have a flexible and collaborative case development platform, allowing us to modify the scenarios based on test user feedback. Twine’s integration with multimedia elements was crucial for reflecting complex clinical scenarios. The platform’s compatibility with various foundational technologies ensured accessibility across multiple devices—a vital feature in resource-constrained environments. This adaptability is reflected in our impressive System Usability Scale score of 74.6 (“Excellent”), affirming the tool’s intuitive and user-friendly design, thereby enhancing the educational experience. With this SUS score our application was verified to be easy to navigate, so that the evaluation of the users’ performance can be mainly attributed to their medical knowledge and clinical reasoning. This is an important, yet often neglected, aspect of modern educational applications, where the technical aspects of the application itself can potentially affect academic performance.

Our framework’s scalability is another noteworthy strength. Relying on foundational web technologies like HTML and JavaScript guarantees long-term viability and adaptability across diverse medical specialties and geographic contexts. Additionally, Twine’s potential for procedural generation of scenarios can introduce unique and diverse learning experiences, exposing trainees to a wide array of clinical presentations. ^16^ The introduction of elements such as achievements or progress tracking in future iterations could further enhance learner engagement and motivation, potentially improving knowledge retention and classroom dynamics.

The framework does also have limitations, including limited server interactivity and analytics capabilities, along with a reliance on third-party data sharing integrations, which may pose privacy concerns. Compared to platforms like DecisionSim and OpenLabyrinth, our approach emphasizes cost-efficiency and customization but provides fewer advanced features. Nonetheless, its adaptability and minimal infrastructure requirements make it invaluable for institutions or individuals constrained by budgets and the lack of technical capacity.

Our use of AI-generated images via DALL-E^17^ to create culturally specific visuals free from copyright issues marked a significant enhancement. AI-generated images were selected as an option for their ease of adaptability, lack of copyright liability and time efficiency. The authors want to acknowledge the concerns that arise given that these models were trained on artistic works without permission or remuneration and suggest that future and/or larger projects take this into account.

Addressing AI bias was also critical, as initial outputs did not reflect the Haitian context accurately. Even though “Haiti” - where the White race represents a minority of the population - was mentioned in the prompt, doctors were shown in the generated images to be White more often than not. This highlights the need for ongoing ethical vigilance when employing AI applications, ensuring educational content is both representative and unbiased.

In conclusion, the GMENEcho framework addressed a critical need in cardiology training tools, particularly in resource-limited settings. By prioritizing accessibility, rapid customization, and cost-effectiveness, our model offers a flexible template for adoption in rapid prototyping and small-scale projects.

## Data Availability

All data produced in the present study are available upon reasonable request to the authors

https://github.com/Tsaftaridis/Virtual_echo

## Ethics and Oversight

The Ethics Committee of the University Hospital of La Paix gave ethical approval for this work. Yale University’s institutional review board (ID: 2000034601) gave ethical approval for this work. Compliance with both local regulations and international ethical standards was ensured. Informed consent was obtained from all participants, who were assured of their right to withdraw from the study at any time without penalty.

## Data Availability Statement

All data produced in the present study are available upon reasonable request to the authors.

## 5 Supplemental Information

The Shapiro-Wilk Test Results for Normality validates the appropriateness of the parametric tests utilized by confirming a normal distribution of the differences between the pre- and posttests. Completion rates were high, with 12/16 completing the pretest and 14/16 completing the posttest.

